# Home Use of a Wireless Intracortical Brain-Computer Interface by Individuals With Tetraplegia

**DOI:** 10.1101/2019.12.27.19015727

**Authors:** John D. Simeral, Thomas Hosman, Jad Saab, Sharlene N. Flesher, Marco Vilela, Brian Franco, Jessica Kelemen, David M. Brandman, John G. Ciancibello, Paymon G. Rezaii, David M. Rosler, Krishna V. Shenoy, Jaimie M. Henderson, Arto V. Nurmikko, Leigh R. Hochberg

## Abstract

Individuals with neurological disease or injury such as amyotrophic lateral sclerosis, spinal cord injury or stroke may become tetraplegic, unable to speak or even locked-in. For people with these conditions, current assistive technologies are often ineffective. Brain-computer interfaces are being developed to enhance independence and restore communication in the absence of physical movement. Over the past decade, individuals with tetraplegia have achieved rapid on-screen typing and point-and-click control of tablet apps using intracortical brain-computer interfaces (iBCIs) that decode intended arm and hand movements from neural signals recorded by implanted microelectrode arrays. However, cables used to convey neural signals from the brain tether participants to amplifiers and decoding computers and require expert oversight during use, severely limiting when and where iBCIs could be available for use. Here, we demonstrate the first human use of a wireless broadband iBCI. Based on a prototype system previously used in pre-clinical research, we replaced the external cables of a 192-electrode iBCI with wireless transmitters and achieved high-resolution recording and decoding of broadband field potentials and spiking activity from people with paralysis. Two participants in an ongoing pilot clinical trial performed on-screen item selection tasks to assess iBCI-enabled cursor control. Communication bitrates were equivalent between cabled and wireless configurations. Participants also used the wireless iBCI to control a standard commercial tablet computer to browse the web and use several mobile applications. Within-day comparison of cabled and wireless interfaces evaluated bit error rate, packet loss, and the recovery of spike rates and spike waveforms from the recorded neural signals. In a representative use case, the wireless system recorded intracortical signals from two arrays in one participant continuously through a 24-hour period at home. Wireless multi-electrode recording of broadband neural signals over extended periods introduces a valuable tool for human neuroscience research and is an important step toward practical deployment of iBCI technology for independent use by individuals with paralysis. On-demand access to high-performance iBCI technology in the home promises to enhance independence and restore communication and mobility for individuals with severe motor impairment.

## I. INTRODUCTION

**N**EUROLOGICAL disease or injury such as amyotrophic lateral sclerosis, stroke and cervical spinal cord injury can result in tetraplegia, loss of speech or locked-in syndrome. Brain-computer interfaces (BCIs) are being developed to restore communication and motor function for individuals living with profound motor disability. Motor BCIs aim to provide access to assistive devices by decoding user commands from electroencephalography (EEG), electrocorticography (ECoG), or intracortical signals. In ongoing clinical research, high-performance intracortical BCIs (iBCIs) are being developed to infer a user’s movement intentions [1]–[8] or speech [9], [10] from neural activity recorded from one or more microelectrode arrays implanted in motor areas of cortex. By imagining natural hand, finger and arm movements, trial participants with paralysis have achieved reach-and grasp with robotic and prosthetic limbs [2], [3], [11] and their own reanimated limb [6], [12], and have demonstrated reliable cursor control for tablet use [8] and on-screen typing for communication [4], [7], [13]. Building on steady advances in point-and-click accuracy, speed [4], [7], [13]–[16] and consistency [17]–[21], iBCI trial participants at home have achieved average on-screen point-to-select typing rates over 37 correct characters per minute maintained over days and weeks [4], [7].

While this progress is promising, one practical limitation of current iBCIs is their reliance on recording cables that link an implanted array’s head-mounted titanium connector (“pedestal”) to the signal processing and decoding computers. Enabling iBCIs for long-term recording and independent mobile use at home without technical supervision will require wireless acquisition of intracortical signals to eliminate tethering cables to the head. However, wireless recording for iBCI has yet to be demonstrated in people, in part because translating the proven cabled system (16 bits per sample at 30 kS/s for each of 96 electrodes per implanted array) to wireless form presents significant engineering challenges. Although wireless broadband signal acquisition might not be a design requirement for some targeted iBCI applications [22], it allows for investigation of novel iBCI algorithms spanning the full spectrum of neural activity while supporting a wide range of fundamental electrophysiological research during untethered use. With this motivation and the goal of enabling continuous, independent use of an iBCI, previous work from our team created a compact, power efficient neurosensor that digitizes and wirelessly transmits broadband cortical activity from a 96-channel chronically implanted microelectrode array [23]–[26]. A battery-powered pedestal-mounted form of that transmitter was demonstrated in animals during open-loop tasks and free home-cage behavior [23], [24]. A fully implanted inductively-charged version was developed and tested *in-vivo* in non-human primates [25], [26] and is on a translational path for human use. Both the external and fully implanted devices were designed with a relatively high sample rate (20 kS/s per electrode, 12 bits/sample) to support both fundamental human neuroscience research and investigational signal processing and decoding methods for high-performance iBCI systems.

Here we report translation of a wireless broadband intracortical BCI system to human use and evaluate its performance during use at home by two participants in the BrainGate pilot clinical trial. The external transmitter underwent commercial manufacturing and preclinical safety testing in preparation for human investigational use. Transmission frequency was configurable such that neural activity from two 96-channel intracortical arrays could be recorded simultaneously without interference. Two transmitters and associated commercial receiver hardware were integrated into the iBCI real-time signal processing system. Both participants then used the BrainGate system to complete a series of cursor-based point-and-dwell assessment tasks [1], [7], [27] to quantify iBCI performance in cabled and wireless configurations. We demonstrate that the wireless signals could be decoded with sufficient quality and reliability to support spontaneous user-paced point-and-click use of a tablet computer as reported for previous participants using a cable connection [8]. To verify wireless function in one representative iBCI use scenario, one participant completed dual-array wireless recording over 24 hours of daily activity, rest and sleep. Finally, we examined the underlying signal quality and digital sample integrity in the wireless data streams relative to the cabled system in both benchtop and in-home evaluations.

## II. Methods

### A. Participants

Study participants T5 and T10 were previously enrolled in the pilot clinical trial of the BrainGate Neural Interface System (http://www.clinicaltrials.gov/ct2/show/NCT00912041). At the time of this study, T5 was a 63-year-old man with a C4 AIS-C spinal cord injury (SCI) resulting in tetraplegia. T10 was a 35-year-old man with a C4 AIS-A SCI resulting in tetraplegia.

All activities, including use of the wireless devices, were permitted by the US Food and Drug Administration (Investigational Device Exemption #G090003) and the Institutional Review Boards of Partners Healthcare/ Massachusetts General Hospital, Providence VA Medical Center, Stanford University and Brown University.

For each participant, intracortical neural activity was recorded from two 96-channel planar silicon microelectrode arrays (4 mm x 4 mm, 1.5 mm electrode length, platinum tips; Blackrock Microsystems, Salt Lake City, UT) placed in the left (dominant) precentral gyrus, except participant T10 whose second array was placed in the middle frontal gyrus.

### B. Standard Cabled iBCI System

The cabled BrainGate iBCI used commercial hardware and software (Blackrock Microsystems) to acquire and record neural signals. This system included a NeuroPort Patient Cable connecting each percutaneous head-mounted pedestal to a Front End Amplifier which applied a hardware filter (0.3 Hz – 7.5 kHz) and digitized signals on each of 96 microelectrodes (30 kS/s, 16 bits per sample). The continuous serial stream of digital samples was relayed over fiber optic cable to a Neural Signal Processor (NSP) where they were timestamped and sent out as UDP packets on a private local area network (LAN). These “raw” data were stored (without software filtering or down-sampling) by Blackrock’s Central Suite software for offline analysis. Because participants in this study each had two arrays, the home iBCI included two parallel 96-channel acquisition systems (Fig. 1, top) that were time-locked by a sync cable linking the two NSPs.

**Fig. 1.**
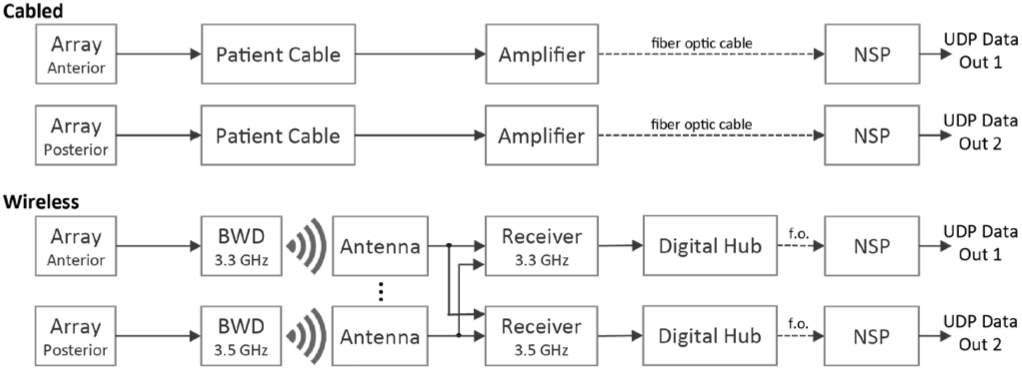
Components of the cabled and wireless systems for dual-array recording. Pathways for neural signal acquisition differed as shown, but NSPs and all downstream file recording, signal processing and decoding hardware and software were the same for both systems.

The raw data packets were also delivered in real-time to downstream Assistive Technology computers for signal processing, neural feature extraction and decoding. The Assistive Technology system included a Super Logics x86 computer (Natick, MA) running the xPC Target real-time operating system (The Mathworks, Natick, MA) and custom signal processing and decoding algorithms running in Simulink. Decoded cursor commands were sent from Simulink to a Dell laptop (T5) or a Microsoft Surface tablet (T10) over wired Ethernet, wireless Ethernet (through a 5GHz WiFi router), or Bluetooth. Bluetooth communication employed our previously developed circuit board dongle based on BlueSMiRF technology (SparkFun Electronics, Boulder, CO) that plugged into the xPC Target computer and presented BrainGate decoder outputs as standard wireless mouse point and click commands [8], [28].

### C. Pedestal-Mounted Wireless Transmitter (BWD)

The neurosensor deployed here (Fig. 2a, b) was designed from the outset for translation to human intracortical recording and in-home BCI applications. Design criteria included a high digital sampling rate to capture neuronal spiking activity, data acquisition and transmission over at least 24 hours for use throughout the day, and lightweight mating to the percutaneous pedestal that is currently the only interface available for chronic intracortical microelectrode recording in people (research only). Details of the design and characterization of the wireless transmitter have been reported previously for external [23], [24] and fully implanted [25], [26] forms. The external device was licensed to Blackrock Microsystems who made minor design refinements and manufactured it as the “Brown Wireless Device” (BWD). The BWD applies a hardware filter (1 Hz to 7.8 kHz), digitizes neural activity (20 kS/sec, 12 bits per sample) on each electrode, applies Manchester encoding and transmits it to nearby antennas using a custom low-power protocol. The Manchester encoding provides self-clocking, reduces link error rate and enhances reliability. In the digital data stream, a 24-bit “sync word” separates each 50 μs “frame” of data containing one 12-bit sample from each electrode. BWDs use a non-rechargeable medical-grade battery (SAFT LS14250, 3.6 V, 1000 mAh). Pre-clinical device safety assessments were completed prior to use in the BrainGate trial.

**Fig. 2.**
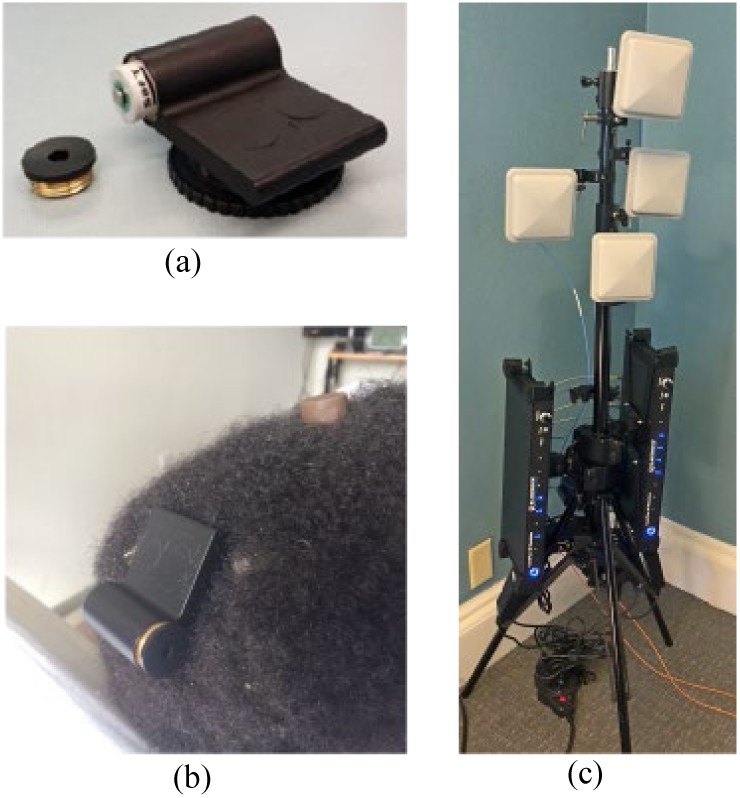
Some components of the wireless system. (a) BWD transmitter (5.2 mm x 4.4 mm) showing battery compartment. Turn-screw disc is used to attach the device onto a percutaneous pedestal. (b) The BWD connected to T10’s posterior pedestal (the anterior pedestal is covered by a protective cap). (c) A two-frequency wireless receiver system in a four-antenna SIMO configuration as deployed for T10. The output optical fibers (orange) connect to downstream NSPs.

### D. Wireless iBCI System

In the wireless recording system (Fig. 1, bottom), each NeuroPort Patient Cable and Front End Amplifier was replaced by four components (Blackrock Microsystems): a BWD, one or more polarized planar antennas (5” x 5”, 3GHz – 4 GHz reception, PA-333810-NF), a Wireless Receiver (PN9323) and a Digital Hub (PN6973). The transmitter was designed as a low-power, short-range (meters) broadband device operating in a low-use portion of the microwave communications spectrum. Each BWD digitized neural activity from one array and transmitted it at 3.3 GHz or 3.5 GHz (configured at time of manufacture) to the antennas. The corresponding receiver was manually tuned to the appropriate frequency. Each Digital Hub relayed the digital data stream to its respective NSP over fiber optic cable. The NSP, file recording system, and downstream Assistive Technology hardware and software were unchanged between the wired and wireless configurations.

The Wireless Receiver supported up to 8 antenna inputs in a Switched Diversity Single-Input Multiple Output (SIMO) architecture for improved reception. SIMO provides robust recovery of a single communication link (one BWD’s continuous transmission) by allowing multiple antennas (e.g., in a home environment) where the signal can be retrieved (i.e., multiple outputs of the communication link). The receiver considered a data frame to be valid on any antenna input when the expected terminating sync word was detected (“signal lock”) which was indicated by a blue LED on the front panel. The sample data in each frame did not include error checking, so it was possible for bit errors to occur in the data even when the sync word was detected. SIMO was used to provide a degree of data checking: when two or more receiver channels reported lock, the data output by the receiver was taken from a channel whose contents exactly matched those of another locked channel.

After preliminary wireless reception testing in the home of T10, we elected to reduce the physical footprint of the wireless system by using four (rather than 8) antennas mounted above and/or behind the user, 1-2 m from the pedestal-mounted transmitters. A tripod stand behind his bed allowed four antennas to be positioned near the head and adjusted to achieve effective angle and polarization orientations (Fig. 2c). For T5, two antennas were positioned on tables near his wheelchair and two at the ceiling slightly behind the wheelchair. During setup for each recording session, we ensured that LEDs on each receiver consistently indicated signal “lock” for each antenna input but did not attempt to further optimize the placement of antennas. As with the wired system, dual-array wireless recording required two parallel sets of equipment, except that the SMA cable from each antenna was split and connected to both wireless receivers. This avoided the need for duplicate antennas at the cost of splitting the signal power between the receivers.

To establish compatibility with the downstream data recording system used for this study, the wireless hardware (Wireless Receiver / Digital Hub) upsampled the BWD data stream from 20 kS/s to 30kS/s (via sample-and-hold) and from 12 bits per sample to 16 bits per sample (four-bit up-shift). The upsampled wireless data was then stored in the standard ns5 file format and processed through the identical signal processing software as the cabled data. We applied a variety of analyses to assess the effect of these sample-level transformations on recorded neural signal content and iBCI decoder function.

### E. Overview of Participant Research Sessions

Prior to the first study session with T10, we performed an in-home spectral sweep for potential radio frequency (RF) interference with a microwave spectrum analyzer (Anritsu). No detectable sources of interference in the 3.0 – 4.0 GHz band used by the BWD were found, despite numerous in-home appliances, medical devices, and a nearby line-of-sight commercial radio tower.

Participant sessions in this study included T10 trial days (post-implant day) 307, 349, 350, 355, 361 and T5 trial days 560, 572, 588. Research sessions took place in participants’ homes with oversight by each site’s trained clinical neurotechnology research assistant (CNRA). T5 completed study activities seated in a wheelchair in his living room. For health reasons unrelated to the neural interface system, T10 completed all sessions while reclined in bed. Each participant completed two sessions in which cursor kinematics were controlled through decoded neural signals recorded in both the wired and wireless configurations. Each participant also completed a third session in which they used decoded point-and-click cursor control to navigate consumer software and web applications on a tablet computer. Sessions lasted approximately 3 hours, interrupted as needed by the participant’s regular nursing and personal care activities. T10 also completed an overnight recording session spanning 24 continuous hours.

### F. Neural Decoding Methods

For iBCI cursor tasks, a Kalman filter decoder and a linear discriminant classifier were used to estimate continuous 2-D cursor velocity and click state, respectively. After upsampling in the wireless path (described above), data processing and decoding algorithms were identical for cabled and wireless conditions as described next. The signal processing and decoding algorithms used here have been described in detail previously [4]. Briefly, signals were downsampled to 15 kS/s followed by common average referencing. For decoding, two neural features were then computed for each electrode: spike rates and spike power. Spike rates were computed from thresholded spikes. Spike power was estimated as the power in the high-frequency local field potential (HF-LFP). For both features, a band pass filter was first applied (250Hz - 5 kHz, 8th order IIR Butterworth). Spike rates were then computed by re-filtering the band-passed data in reverse to provide net zero-phase shift [29] before applying a spike threshold at -3.5 standard deviations (computed for each channel from prior within-day blocks of data). Spike power was computed by squaring and averaging the band-passed signal. Spike power and thresholded spike rate features were z-score normalized using each feature’s mean value and standard deviation calculated from a previous data block. All features, velocity, and state estimates were updated every 20 ms.

### G. Decoded Cursor Control in Cabled versus Wireless iBCI

During participant sessions, cabled versus wireless cursor performance was evaluated using an A/B block design in which decoder calibration and several back-to-back Grid Task cursor assessments were completed in the cabled configuration (A) followed by the same sequence using the wireless system (B). For each condition, participants first completed a 3-minute long cursor-based closed-loop calibration task [27] which automatically computed coefficients of a 2-dimensional Kalman filter. For tablet control sessions, a discrete “click” state classifier was also calibrated. Participants commanded cursor movement and dwell using attempted hand (T10) or finger (T5) movements. Clicks were commanded using attempted hand grasp (T10) or attempted thumb tap (T5). After decoder calibration, several back-to-back Grid Task assessments were completed [1], [7], [27] to quantify iBCI-enabled closed-loop point-and-select performance. Switching between cabled and wireless configurations involved powering down the iBCI equipment, removing the NeuroPort Patient Cables, connecting BWDs to the two pedestals, replacing the Front End Amplifier fiber-optic cable into the NSP with the fiber optic cable originating at the Digital Hub, and restarting the hardware and software. Decoder coefficients were randomly re-initiated prior to the automatic calibration step that started each series. In another series on the same or different day, the entire sequence was reversed to achieve a B/A block design (wireless first) to balance possible order effects.

### H. Grid Task Assessment of iBCI Cursor Control

Decoder performance in wired and wireless conditions was assessed on a series of Grid Tasks (2 minutes each) in which participants moved the cursor to the indicated target and then dwelled on the target to select it. The Grid Task consisted of a *6* x *6* grid of adjacent squares in which all points were selectable as described previously [1], [7], [30]. Dwelling on any target for 1 second selected it (resulting in a correct or error selection) after which the next target was shown. Selections continued uninterrupted until the Grid Task ended. If no target was selected within 8 seconds then a timeout resulted in an error trial and the next target was presented. The Grid Task was repeated several times with brief pauses in between. Task performance was assessed for each 2-minute task in terms of Percent Correct target acquisitions (hit rate) and Bitrate quantifying information transfer (bits per second) as reported previously for this task [31]. Metrics of cursor control included Trial Duration (average time to acquire targets including successful dwell time and the timeout duration for error trials, in seconds), Path Efficiency (ratio of actual trajectory length from starting point to the target relative to the straight line path), and Angle Error (the mean angle between instantaneous trajectory and the direct-to-target trajectory, in degrees).

### I. Wireless iBCI Use of a Tablet Computer

To evaluate whether the wireless neural interface system could enable individuals with paralysis to achieve reliable point-and-click control of a computer in their homes, T5 and T10 used the wireless iBCI for point-and-click control of an unmodified consumer mobile device (Microsoft Surface Pro tablet) for web browsing, use of consumer applications, and typing communication. Cursor control was enabled using our standard automated decoder calibration, then CNRAs placed the tablet comfortably in front of the participant and routed the iBCI decoded cursor and click commands to the tablet [8]. Participants used the tablets in Microsoft Windows 10 “tablet” mode to navigate the desktop and to start and use different programs of interest to them. Additionally, T5 was asked to complete several blocks of typing using Windows Notepad to provide estimates of typing performance. Although the native onscreen keyboard offered word prediction, T5 avoided these and selected all letters, capitalization (“shift” function), spaces and punctuation explicitly. For each typing block we computed correct characters per minute (ccpm) as the number of appropriate character selections including spaces and punctuation (excluding characters resulting in misspelling, “shift” selected for capital letters, and delete or backspace key selections) and dividing by the elapsed time. Detailed analysis of application use and typing rates were performed through offline analysis of screen capture recordings.

### J. 24-Hour Wireless Recording

T10 completed an overnight in-home study to collect nearly continuous intracortical data over a 24-hour period. The study began with a typical CNRA-administered BrainGate research session in which data were collected during point-and-click cursor calibration and performance assessment. A similar research epoch was completed the next day to end the 24-hour period. Between these administered research periods, the wireless system was configured to continuously record and store raw neural data for offline analysis. For this first-ever overnight recording, two CNRAs maintained continuous monitoring on a rotating schedule. During this time, T10 remained in bed for reasons independent of this study, but he engaged in his typical daily activities such as eating, watching television, conversing by phone, direct family interactions, and sleeping. He received his regular nursing care including rotations in bed every few hours. Because RF pathway obstructions were anticipated as caregivers stood or moved between the transmitter and antennas, and because body rotations could alter orientation of the two transmitters relative to each antenna, a second pole of 4 antennas was added for this overnight study (increasing the number of antennas from 4 to 8). Based on laboratory testing, the battery in each transmitter was proactively replaced every 10 hours to avoid any possibility of unanticipated signal loss due to battery drain. (The wireless transmitter was designed for 48 hours of continuous use per battery [23]; after this study, a correction to the BWD restored it to 48+ hours). A video camera captured snapshots of the room once per minute (with participant permission) to document activity. File recording software (Cerebus Central File Storage App, Blackrock Microsystems) automatically split neural data into contiguous files (with no sample loss) to limit the size of individual files.

Offline, integrity of the recorded data was analyzed in 5-minute segments contiguously across the 24-hour period (288 segments for each array). For visualization, the spectral content was computed from 0.5 Hz to 100 Hz in each segment. A “disrupted” segment was one with missing recorded data (e.g., brief stops between CNRA sessions and overnight recording), dropped packets, or pervasive noise (identified as segments in which the summed spectral power from 40 Hz to 100 Hz exceeded the 90^th^ percentile of the entire 24 hour period).

### K. Comparing Cabled and Wireless Signal Fidelity

We conducted a series of A/B experiments to more directly compare the fidelity of broadband recordings using the standard 96-channel NeuroPort Patient Cable “reference system” and the 96-channel wireless system. We compared noise, bandpass filtered signals, sorted spike waveforms and unit firing rates. These tests were performed in the lab and with participants at home.

#### Bench Measurements

An initial baseline comparison of signal fidelity was performed using a single NeuroPort Patient Cable or BWD transmitter to record well-defined signals from a Neural Signal Simulator (NSS, Blackrock Microsystems). Four antennas were placed within three meters and direct line-of-sight of the wireless transmitter to reduce the possibility of wireless data drops that could confound these baseline evaluations. The neural signal simulator provided continuous, well-defined patterns of voltages simulating slow local field potential oscillations (1 Hz, 3 Hz, 9 Hz and 20 Hz), three different repeating single-unit action potential waveforms on each channel, and baseline noise. Several blocks of broadband data (∼5 minutes each) were recorded in consecutive wired and wireless configurations and then analyzed offline. Because the signal simulator generated deterministic spiking patterns, it was possible to precisely align and compare the continuous raw recorded signals and spikes (waveforms) across conditions. To objectively compare spiking units without operator bias [32], we first applied automated spike-sorting to each data set using a previously reported algorithm [33] and compared the resulting sorted unit waveform shapes, spike rates, and SNR measures.

#### Intracortical Signals

These head-to-head comparisons evaluated data recorded with T5 and T10 during the closed-loop point-and-click Grid Tasks recorded in multiple interleaved cabled (A) and wireless (B) data blocks within each assessment day. A/B studies were repeated in a different order on separate days with both participants. As with bench measurements, comparisons of recorded spiking activity were made after automated spike sorting.

To compare signal amplitudes across wired and wireless conditions, we applied an empirical gain adjustment to match absolute voltages in the recorded signals. Gain stages in the commercial wireless hardware were designed to achieve the same theoretical total gain as the wired hardware. However, the precise cumulative gain in each recording system is sensitive to normal variability in device fabrication parameters. These resulted in a small but measurable and consistent gain difference between wired and wireless recorded signals. We calibrated absolute voltages by finding the gain multiplier that minimized differences in the NSS simulated spike waveforms recorded in the wired and wireless bench tests. Based on these empirical measurements, we applied a +2.97% amplitude adjustment to the wireless data for these comparisons.

### L. Digital Sample Integrity

Transmission data loss is a concern for any wireless system. Here, wireless data loss occurred whenever the receiver did not find the expected sync word on any input channel. When this occurred, the current samples from all 96 electrodes were invalid and the previous valid frame was inserted into the data stream. This frame error was detectable in the recorded data as a back-to-back repeat of the previous 96-channel data sample. We quantified the number of frame errors in the received 20 kS/s wireless data by examining the 30 kS/s data recordings and accounting for the expected back-to-back sample repeats from the upsampling process. The practical effect of wireless data loss depends not just on the number frame errors but on their distribution. Individual dropped frames (one 50 μs sample from each electrode) distributed sparsely throughout the data stream should be less detrimental to iBCI decoding than contiguous data loss [22]. We quantified the prevalence of longer intervals of data loss using the standard Severely Errored Seconds (SES) metric that reports the number of non-overlapping 1-second periods in which the proportion of data errors exceeds a predetermined threshold. Here, a commonly used threshold of 50% found all 1-second periods in which frame errors corrupted at least 500 ms of the data.

We interrogated the wired and wireless 30 kS/s recorded data for all instances of voltage changes exceeding a pre-determined biologically plausible slew rate (500 μV per 33 μs sample). These included bit-flips within single samples resulting in a large voltage excursion that returned to baseline in the following sample (or next sample after that in the case of upsample repeats), and large amplitude noise events extending over many samples before returning to baseline. We examined the degree to which each event was isolated to individual channels or observed simultaneously across many electrodes of the recording array.

## III. Results

### A. Translation of the Wireless Sensor for Human Studies

The commercialized BWD underwent pre-clinical device testing in accordance with U.S. Food and Drug Administration (FDA) Good Laboratory Practice (GLP) guidelines. BWD testing verified safe levels of charge transfer; reliable sterilization (ANSI/AAM/ISO 10993-7-2008); immunity to repeated direct and indirect electrostatic discharge (ESD) up to +/-6kV and +/-8kV, respectively (IEC 61000-4-2). Radio frequency (RF) transmission power measured well under 1 mW, below the specific absorption rate (SAR) exclusion threshold of 16 mW (FCC 447498 D01 General RF Exposure Guidance). The transmitter was operated in high density RF environments ranging from a major hospital to a metropolitan city center with no interference from such telecommunications-rich environments. The FCC granted an Experimental License to operate the BWD in the 3.3 GHz – 3.8 GHz transmission band in geographical regions related to the BrainGate trial.

### B. Assessment of Wireless Point and Click Cursor Control

We evaluated if the cabled and wireless iBCI configurations enabled equivalent kinematic decoding and closed-loop cursor performance by BrainGate trial participants in their homes. On each of two days, participants successfully achieved cursor control using an automated calibration procedure which generated the Kalman decoder coefficients used in the subsequent Grid Task assessment blocks. Across two sessions, T10 completed 172 trials over 6 blocks in the cabled configuration and 167 trials over 6 blocks in the wireless configuration. Participant T5 completed two sessions as well totaling 922 trials over 20 blocks in the cabled configuration and 1,023 trials over 24 blocks in the wireless configuration.

The percentage of successful target selection was not significantly different between wired and wireless systems (Wilcoxon rank sum > 0.05) for either participant (Fig 3a). T5 median success rate was 96.2% versus 97.9% in the cabled and wireless conditions, respectively. T10 achieved 91.6% and 94.8% in cabled and wireless configurations. Bitrates in the Grid Tasks were also not significantly different (Fig. 3b) between cabled and wireless for either participant (Wilcoxon rank sum > 0.05; T5: 1.58 bps cabled and 1.77 bps wireless; T10: 1.47 bps for both systems).

**Fig. 3.**
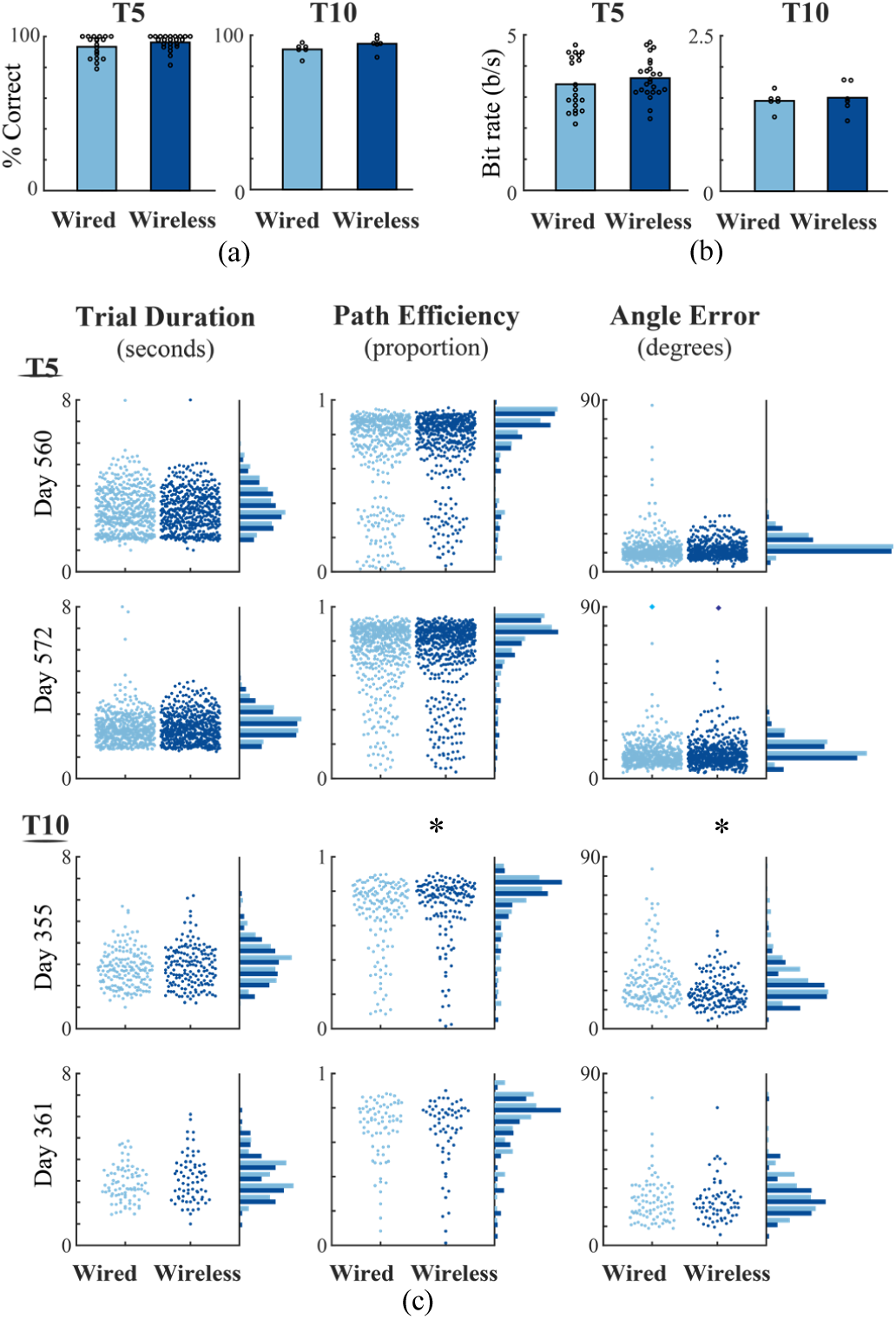
Metrics comparing closed-loop cursor control in wired (light blue, left half of each figure) and wireless (dark blue) configurations. (a) Median target acquisition rates in wired and wireless conditions. Circles indicate the measure for each iteration of the Grid Task across two sessions for each participant. (b) Bitrates in wired and wireless conditions (one measure for each grid task across two sessions for each participant). (c) Three metrics of cursor control over two days for each participant. Each point shows the metric computed for an individual trial (one target acquisition). Points are spread on each X-axis to reveal individual trials. Histograms on the right of each plot summarize wired and wireless performance.

These statistics provided one measure per task iteration. We also quantified cursor control at the resolution of individual trials (Fig. 3c). Across three metrics (trial duration, path efficiency, angle error), no significant differences were observed on either day with T5 or on trial day 361 with T10. Significant differences were observed only for T10 path efficiency and angle error on day 355 (Wilcoxon rank sum < 0.05). On this day, the median measures for both path efficiency (0.74 wired, 0.78 wireless) and angle error (20.7 degrees wired, 18.0 degrees wireless) were better with the wireless system. While the causes of this slight apparent superiority of the wireless system are not clear, the wired condition exhibited more outlier trials with low path efficiency and high angle error on this day.

### C. Wireless iBCI Use of a Tablet Computer

T5 and T10 each used the wireless iBCI to achieve point-and-click control of a standard consumer Microsoft Surface 2-in-1 tablet computer running the Windows 10 operating system. On trial day 307, after completing an autocalibration sequence and several Grid Task assessment blocks, T10 started several widely-used apps in sequence by selecting the corresponding app tile on the Windows start menu or desktop. He used the Microsoft Edge browser, Pandora, the NCAA basketball app, Skype, YouTube, Gmail and the Weather app (Table 1). T5 used these same apps (excluding NCAA) on his trial day 588, including typing on the Windows on-screen keyboard to perform searches (Fig. 4). T5 also typed sufficient spontaneous text in the Windows Notepad to provide a basic measure of typing performance. Over 814 seconds (14.6 minutes), T5 typed 196 correct characters; any characters contributing to misspelling or that T5 subsequently deleted were included in the elapsed time but not in the character count.

**Table 1.** Tablet apps used by T5 during wireless point-and-click control.

| App | Activity |
| --- | --- |
| Pandora | Click to start Pandora in a web browser, select an artist, click the play button, select the "Thumbs Up" icon, click Pause, return to desktop |
| NCAA App | Select app tile on the desktop, select a game "recap" icon, select the game video to play |
| Skype | Select the Skype icon in the start menu, select a contact and start a voice call, talk for 90 s |
| YouTube | Start app from the start menu, select on-screen keyboard icon, type search term, select a video, select "full-screen" icon, click back to desktop |
| Gmail | Click on Inbox icon, select a message and read it |
| Weather App | Click through weather forecasts |

**Fig 4.**
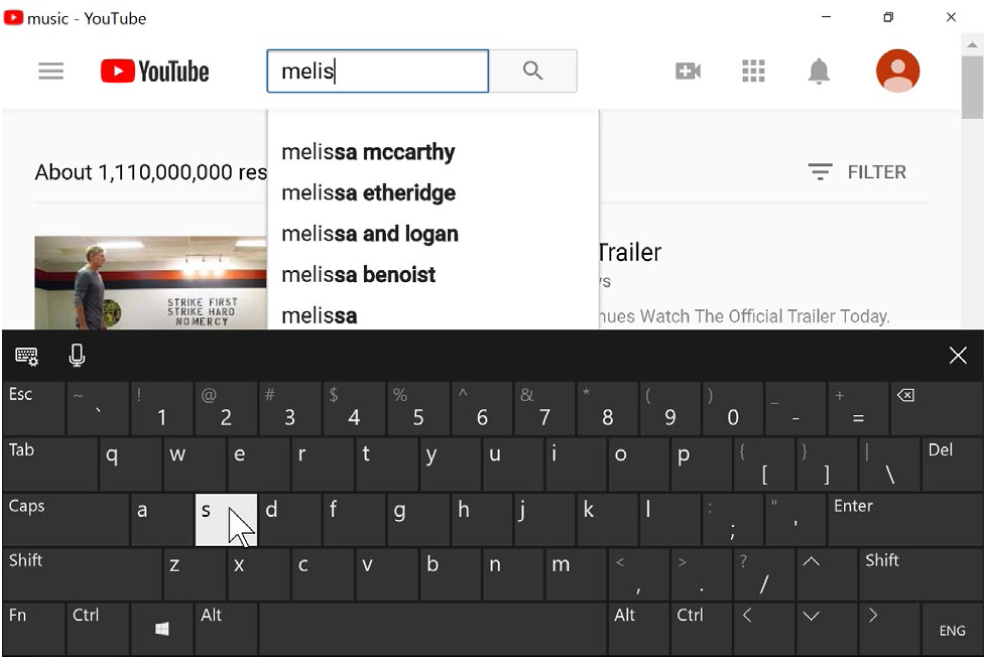
Screen capture of T5’s tablet while he used the iBCI for point-and-click text entry with the built-in on-screen keyboard to search a web site for music by Melissa Etheridge.

### D. Wireless Acquisition During 24 Hours in the Home

In T10’s 24-hour study, synchronized neural data were wirelessly recorded two intracortical arrays from 2:22 pm trial day 349 to 2:22 pm trial day 350. With pre- and post-session activities, total participant engagement time was 26 hours. Each array logged 500 GB of broadband data (1 TB for 2 arrays over 24 hours).

We examined the spectral content of the recorded signals for evidence of transmitted neural activity and/or recording artifacts. Spectral content of the record across the 24 hours was evaluated in contiguous 5 minute segments. Results for the posterior pedestal (recording from the intracortical array in the precentral gyrus) are shown in Figure 5. Prominent LFP activity was observed in bands centered at 10.6 Hz and 19.5 Hz. Clear changes in the spectral power were evident during the night and early morning (e.g., in the ∼20-40 Hz band) corresponding to the period when T10 was observed to be sleeping (approximately 3 am to 9 am).

**Fig 5.**
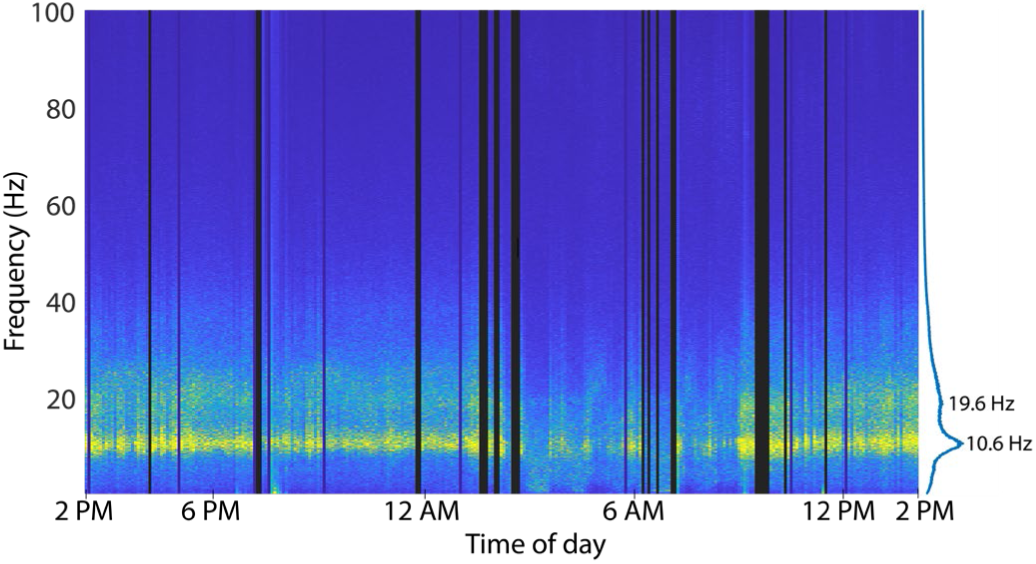
Spectral content of T10 neural data recorded continuously over 24 hours with the wireless system. X-axis indicates wall-clock time. Dark vertical bars reflect periods where data was not recorded (e.g., transmitters removed) or was severely disrupted (high frame loss).

Over the 24 hours (1,440 min.), wireless recording was highly reliable. Wireless recording was uninterrupted throughout most of T10’s activities, including using a head tracking system to type emails and browse the web, having a phone conversation, watching TV, talking, eating, sleeping and more. However, “disruptions” in the form of unrecorded data or noise were observed in 35 of the 288 five-minute analysis segments (175 total disruption minutes). A review of the session logs found that the large majority of data disruptions (100 minutes) occurred when one or more caregivers were attending to T11 including rotating or shifting him in bed, suctioning, and other nursing care (Table 2). During these periods, caregivers worked in close proximity to the bed including standing directly between the transmitters and antennas for several minutes at a time. Data were recorded but exhibited packet loss which was sometimes accompanied by substantial noise when the signal was recovered. In other cases, data flow stopped entirely when transmitters were removed during battery replacement, bathing and changing. Other disruptions were unrelated to the wireless components but rather resulted from technical glitches in the real-time data filing process. Only 15 minutes of data disruption were not readily associated with noted activities; these occurred during sleep between 6:45AM and 7:15 AM.

**Table 2.** Events associated with disrupted wireless signals over 24 hours.

| Total Minutes | Activity |
| --- | --- |
| 100 | Caregivers attending T11 (rotate in bed, suction) |
| 25 | Bathing, dressing (transmitters removed) |
| 15 | Unexplained signal noise during sleep |
| 15 | Glitches in real-time file processing system |
| 10 | Battery changes |
| 10 | File breaks before/after cursor sessions |

### E. Benchtop Comparison of Cabled and Wireless Recordings

To provide a baseline comparison of the two systems, we performed benchtop tests in which well-defined simulated neural signals from the Neural Signal Simulator were recorded through the cabled and wireless pathways. The NSS generated continuous broadband data which superimposed noise, low-frequency oscillations and “action potentials” mimicking three spiking neurons with distinct waveform shapes. After applying a spike-band filter and aligning corresponding simulated spike waveforms, there was a close correspondence between the signals recorded with the two systems (Fig. 6a). We observed low overall noise levels in both conditions with a few microvolts more noise (rms) in the spike band with the wireless system (Fig. 6b; wired noise: 11.2 μV; wireless: 13.5 μV) and 10 μV more noise with the cable in the LFP-filtered band (Fig. 6c; wired: 166 μV; wireless: 156 μV).

**Fig. 6.**
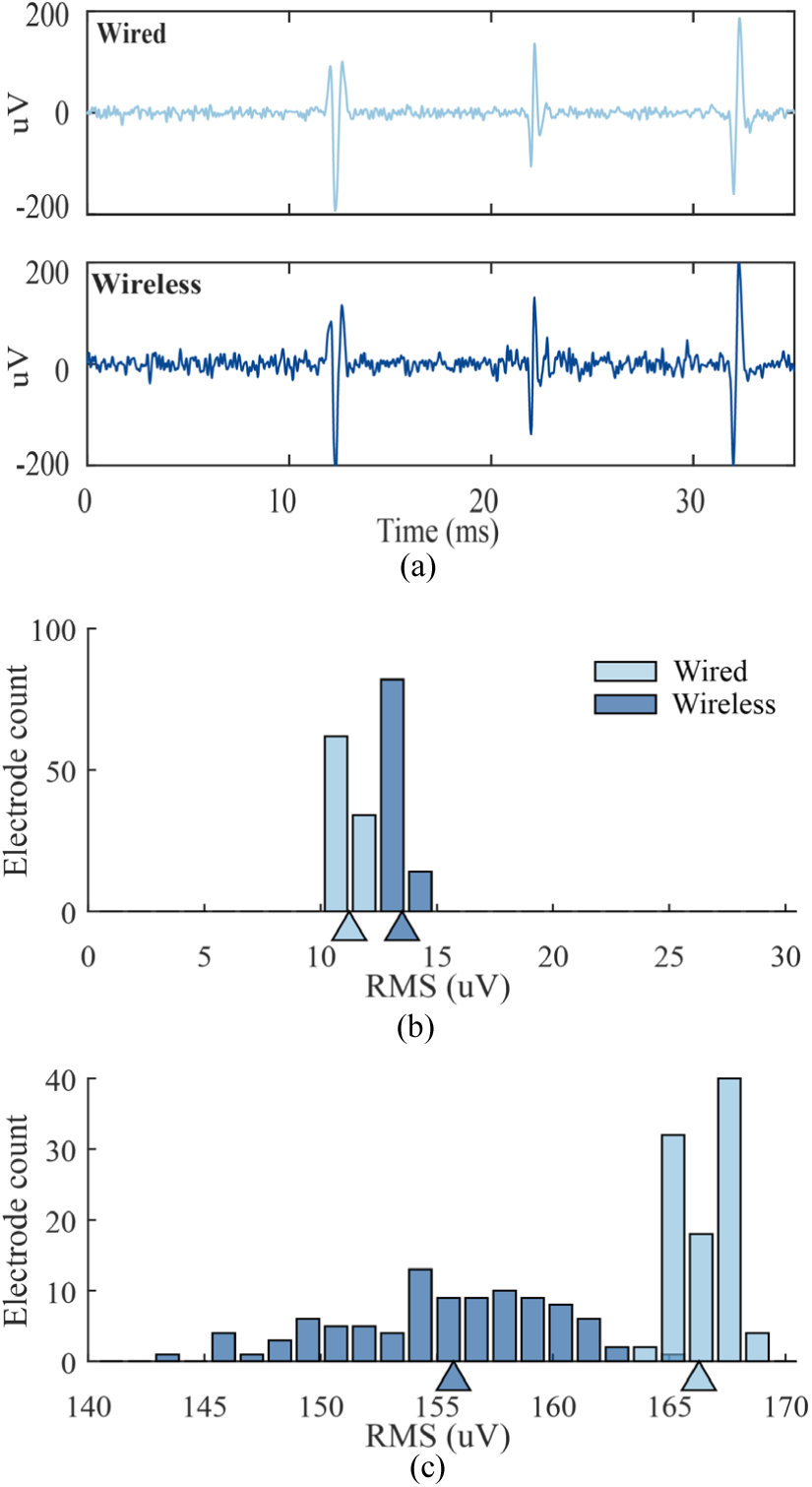
Comparison of wired and wireless recordings of simulated neural signals. **(a)** Waveforms of three different simulated spiking neurons aligned from bandpass-filtered wired and wireless data. **(b)** Distribution of noise across 96 channels filtered for spiking data (250 Hz – 7.5 kHz); baseline noise was measured after spike events were removed by thresholding. Triangles indicate median rms values. **(c)** Distribution of noise across LFP bands (1 Hz – 250 Hz). T10.2018.04.16 002(1)

To evaluate the fidelity of spike recording, we applied an offline automated unsupervised sorting algorithm to recover the waveform shapes and firing rates on all electrodes. The simulator’s three unique waveform shapes were all reliably recovered on all electrodes as indicated by equivalent waveform sorting templates in wired and wireless conditions. The firing rates of all units on all 96 channels were identical (3.6 Hz) and matched the simulated rates, confirming that action potentials were recorded with the requisite fidelity. The equivalence of units recovered from wired and wireless data sets was evident when mean waveforms and their standard deviation were overlaid (Fig. 7).

**Fig. 7.**
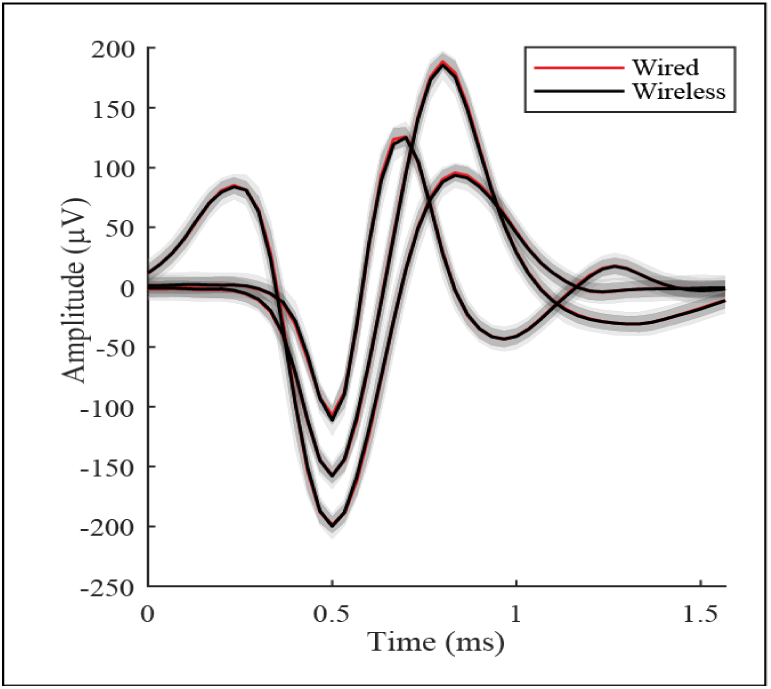
Mean waveforms of the three simulated neurons as extracted from one representative electrode recorded by the cabled (red) and wireless (black) systems. Each unit shows mean (red or black line) +/- s.d. (gray bands) waveform averaged over a random selection of spikes.

### F. Wirelessly Recorded Human Intracortical Signals

In a similar way, we compared human data recorded with wired and wireless systems during all Grid Tasks. The raw (unfiltered), LFP-filtered, and spike-filtered signals were substantially similar across conditions (Fig. 8a). Noise in the spike-filtered intracortical data was slightly lower than observed with the simulator. Consistent with the NSS tests, noise in the spiking band was a few microvolts higher in the wireless condition (Fig. 8b). Spike waveforms were equivalent between the recording modalities as demonstrated by overlaying wired and wireless waveforms for putative single units (Fig. 8c). We extracted waveforms for spiking units on all electrodes, applying the same unsupervised algorithm and parameters to wired and wireless data recorded during Grid Tasks. Comparable single unit waveforms were extracted across most electrodes. However, several units were observed in only one condition or the other. These were usually, but not always, small amplitude units or waveforms that were inconsistently split into multiple units. Thus, the population of extracted units was not identical between wired and wireless data. However, we also observed similar variability between Grid Task blocks recorded with the cable alone. Given the fidelity of recording confirmed in our simulator tests, these discrepancies were most plausibly attributed to nonstationarities in the recorded intracortical signals [17] rather than failures in the wired or wireless systems.

**Fig. 8.**
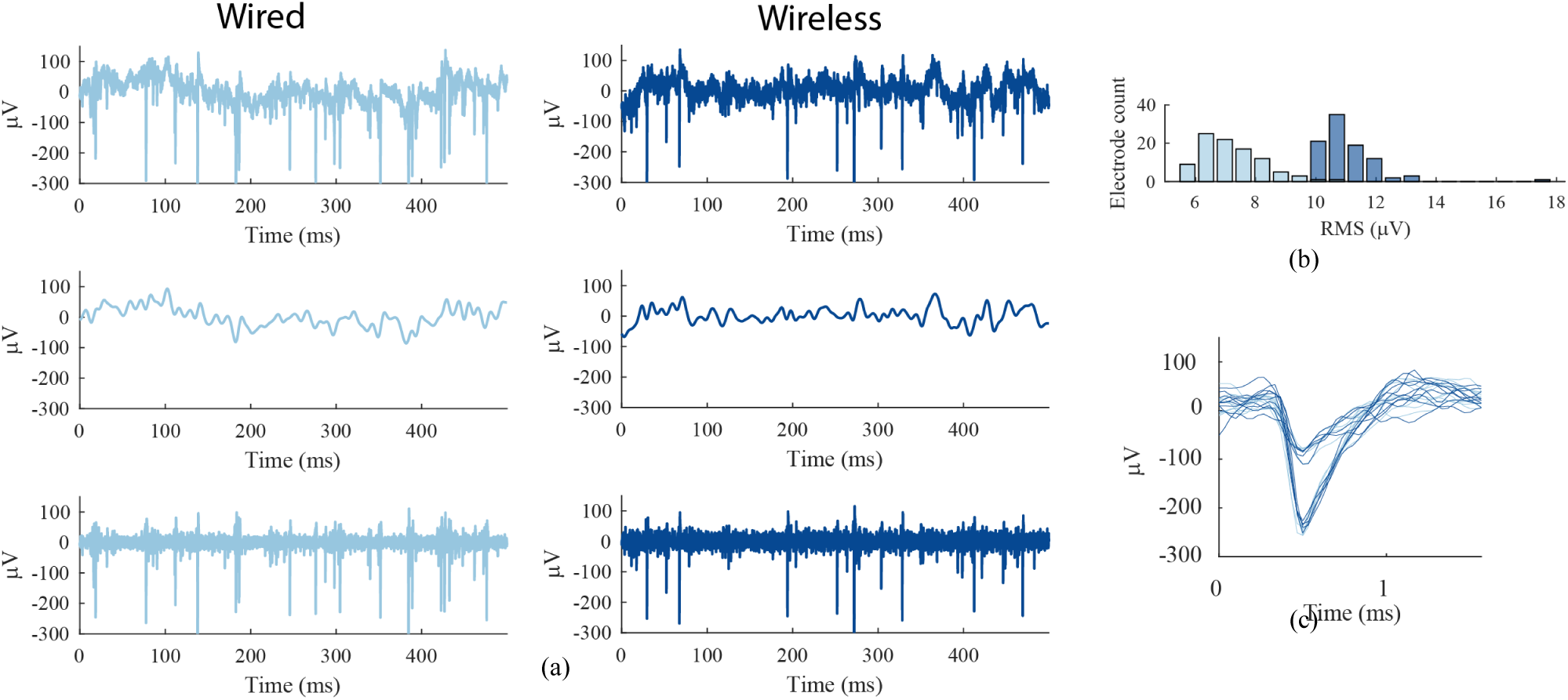
Human intracortical signals recorded in wired and wireless configurations in the home. **(a)** Comparison of recorded neural activity on one electrode (t10.2017.06.05.e24 blocks 6, 7). Top: the “raw” unfiltered neural signal. Middle: low-pass filtered (100 Hz cutoff). Bottom: band pass filtered for spike extraction (250 Hz – 7.5 kHz). (b) Distribution of residual rms amplitudes from all electrodes on one array after band-pass filtering and removing thresholded spikes for wired (light blue) and wireless (dark blue) recordings. (c) Sample waveforms from two units sorted from the same electrode shown in (a) and (b). Light blue (wired) and dark blue (wireless) waveforms show substantial similarity.

### G. Wireless Packet Loss During In-Home iBCI Use

To quantify the incidence of data loss during use of the wireless system, we inspected all raw 30 kS/s data recorded from each participant’s two arrays during all Grid Tasks completed by T5 (24 blocks, 47.9 min) and T10 (6 blocks, 10.3 min) over 2 days each. A “packet drop” occurred when the sync word delimiting a 50 μs wireless data frame containing a single sample from all electrodes could not be recovered by the receiver, resulting in a repeat of all electrodes’ previous digital values being re-inserted into the receiver’s output stream.

The incidence of dropped packets varied across participants and pedestal position (Table 3). For each participant, signal recovery was consistently more successful for one transmitter (anterior 3.3 GHz for T5, posterior 3.5 GHz for T10) than the other. For T5, packet drops were observed during 8 of 24 blocks from the anterior pedestal and during all 24 task blocks from the posterior pedestal; the proportion of dropped packets was overall very low (0.003% of all frames, anterior; 0.4163% of all frames, posterior). No packet drops were ever observed for T10’s posterior pedestal and rarely for the anterior pedestal (fewer than 5 total packet drops per block). In one block, however, T10’s anterior packet drops occurred with a drop rate of 4.69%. We attribute differences in the fraction of dropped packets among blocks to changes in orientation of the participant (transmitters) relative to the antennas. Although participants had limited movement, the four antenna configurations that we used to simplified session setup in the home may not have provided sufficient simultaneous spatial coverage for both directions of transmission from two BWDs.

**Table 3.** Incidence of wireless data packet drops during Grid Tasks. Data for simultaneous anterior (Ant) and posterior (Pos) data streams.

| ID | # Task Blocks | Total Time (Min) | # Blocks with Dropped Packets |  | % of All Data Dropped |  | # of Blocks with SES |  | Total SES Time (sec) |  |
| --- | --- | --- | --- | --- | --- | --- | --- | --- | --- | --- |
|  |  |  | Ant | Pos | Ant | Pos | Ant | Pos | Ant | Pos |
| T5 | 24 | 47.9 | 8 | 24 | 0.003 | 0.416 | 0 | 5 | 0 | 10 |
| T10 | 6 | 10.3 | 6 | 0 | 1.004 | 0 | 0 | 0 | 0 | 0 |

It was noteworthy that during the 2-minute block with high packet drop, T10 nonetheless acquired 44 of 45 targets and recorded his highest bit rate in this study (1.791 bps). This could reflect the fact that, during this block, data from the second transmitter experienced 0 dropped packets. The performance impact of packet drops may also relate to whether they are sufficiently distributed to avoid extended periods of impoverished neural decoding. To quantify this, we computed the number of Severely Errored Seconds (SES), any non-overlapping 1-second period in which half or more of the data samples were lost. Although packet drops occurred, SES events were rarely observed, limited to T5’s posterior data stream in 5 blocks with a cumulative total of 10 seconds. During this study, a total of 6984 wireless data seconds (nearly 2 hours) were recorded during the Grid Task assessments cumulatively across both participants and both arrays; 10 of these exhibited SES.

### H. Bit Level Analysis of Wireless Data

We examined the raw data files for any evidence of bit-level anomalies in the 30kS/s data upsampled from each transmitter. These analyses excluded epochs with signal anomalies attributed to dropped packets (analyzed above). The wireless data exhibited occasional instances of large instantaneous voltage discontinuities. These were biologically implausible voltage differences between consecutive 33 μs samples caused by bit flips among the most significant bits (MSBs) of a digital sample. The corrupt voltage value lasted only one sample (or two samples when an upsample repeat followed) then returned to baseline in a single step. These fast single-sample slew rates were sometimes observed among the lower bits as well, and it seems probable that lower bits were equally prone to flipping but the corresponding small voltage fluctuations would generally have been indistinguishable from baseline activity (and, presumably, functionally inconsequential). The incidence of MSB errors was inconsistent across channels and across blocks. In any given recording, some channels could be recorded error free while others exhibited rare, or more frequent, bit errors. Nonetheless, bit errors were observed in every wireless recording (at a rate of 2.04E-5 to 9.26E-5 per sample throughout the 30 Grid Task blocks). Another form of bit flip persisted for three or more samples and recurred multiple times over a period of tens or hundreds of milliseconds with seemingly valid sample data in between. These periods of recurring MSB errors, which we termed “digital noise”, presented as noisy epochs in the voltage data. Digital noise events were rare in our Grid Task data but more likely in the 24-hour data. In general, we observed a higher incidence of all bit errors including digital noise after periods of packet drops and in association with putative antenna switching events. Bit errors also appeared to be associated with the upsampling logic (see below). Bit errors could have been introduced at various stages of the wireless system such as the BWD analog-to-digital converter, the wireless link, SIMO antenna switching, and the clock domain boundary in the upsampling process. The data collected here did not allow concrete identification of the source(s) of these errors. MSB errors were not observed in the cable-recorded data.

Because MSB errors were identified in our preliminary work, this study applied an algorithm to detect MSB-corrupted samples (briefly, any voltage shift 500 μV or greater within a single sample, i.e., a 15 μV/μs slew rate) and replace each with its preceding good sample on that electrode. In this way, the impact of flip-bit errors was minimized in real-time, prior to feature extraction and decoding, and in offline analyses.

As expected by design, the recorded wireless data contained a large proportion of 33 μs data frames that contained perfect copies of the prior samples on all 96 electrodes. These “upsample repeats” resulted directly from the sample-and-hold logic that upsampled the 20 kS/s BWD data to the NSP 30 kS/s format. Unexpectedly, one or more bits in the upsample repeat frame occasionally differed from their original value. Although these anomalies needed to be accounted for during analysis, they did not otherwise impact wireless performance.

## IV. Discussion

This study demonstrates the first human use of a broadband wireless intracortical brain-computer interface. Two human subjects directed computer cursor movements and click decoded by an iBCI that acquired and wirelessly transmitted (previously prototyped as [24]) broadband neural activity from 192 chronically implanted microelectrodes. Across multiple signal and iBCI performance metrics, the wireless solution proved to be a thorough replacement for the cabled connection currently used in chronic human iBCIs studies and fundamental non-human primate research. Raw neural data were processed into spike rates and LFP power in the spike band which were decoded together to yield precise iBCI cursor control. Using the wireless iBCI, study participants achieved communication bit rates in a Grid Task equivalent to the wired system. Self-paced cursor control enabled both participants to browse the web and complete other tablet activities, and one participant used the wireless interface for free typing. The wireless technology also enabled continuous recording of intracortical broadband field potentials and spiking activity from one participant over 24 hours in his home. Untethered recording of intracortical signals and in-home iBCI use are major steps toward an on-demand iBCI to provide independent digital communication and computer access for people with severe motor impairment.

Evaluation of the raw neural signals and spike waveforms found that signal processing differences in the cabled and wireless pathways had negligible impact on the quality of the recorded neural signals. A small increase in baseline noise with the wireless system was detectable but had no functional impact. The wireless system did exhibit bit errors which we were able to mitigate with real-time algorithms. It proved possible to record broadband intracortical signals in the home without packet drops, as demonstrated by many data blocks in which one or the other transmitter exhibited zero packet drops. Packet drops that did occur were exceedingly rare, indicating the fundamental reliability of the wireless system. Because wireless signal recovery is sensitive to plane of transmitter orientation and distance relative to the position and polarization orientation of the receiving antennas, we anticipate that packet drops could be reduced by more optimal placement of the antennas or by using the full complement of antennas supported by the receivers. Critically, the fidelity of the broadband neural signals and iBCI performance were robust to the low incidence of packet drops that was observed in most blocks. During 24-hour in-home recording we found that volitional head movements and even body rotation by nursing staff generally did not interrupt the wireless signal. However, someone standing or working close to the participant’s head could block transmission entirely, resulting in data loss or “digital noise” during some periods of nursing care. For this study, we elected not to modify users’ homes or wheelchair to accommodate more robustly-placed or permanent antennas. However, these findings motivate further investigation of optimal antenna placement in the home.

### A. Design trade-offs

The BWD system deployed in this study prioritized uncompromising signal fidelity and high electrode count to facilitate both iBCI applications and fundamental human electrophysiology research. However, as the field progresses toward higher and higher electrode counts, technologies for broadband recording become increasingly challenging and alternative, lower-bandwidth signal acquisition approaches are being considered. Design specifications for a wireless targeted BCI multielectrode acquisition system could be simplified by limiting signal acquisition to spiking events (with or without sampled spike waveforms) without broadband data. Although sorted single-unit activity and threshold crossings contain distinct information [34], [35], decoding thresholded multiunit activity rather than sorted units can enable precise iBCI control in monkeys [36]–[38] and in our own BrainGate participants with tetraplegia [4], [7]. Decoders built on multiunit thresholds can also be maintained over long periods [4], [7], [39] and population dynamics may be estimated from multiunit threshold crossings without spike sorting [40]. An analysis of iBCI decoding in monkeys and humans proposes that digital sampling at 1 kHz should be sufficient for iBCI applications [22]. These approaches could reduce the wireless bandwidth by roughly an order of magnitude, thereby simplifying the engineering design constraints for a fully implanted wireless iBCI or, alternatively, freeing bandwidth to record from thousands of electrode contacts [41]–[46].

Despite the broadband design requirement, the BWD design also prioritized low power for long battery life to facilitate users with disability using the system throughout their day. The low power design also translates into fewer inductive charging periods for the fully-implanted version of the BWD (currently in pre-clinical testing). There are several clear trade-offs, however, and comparison to other systems is informative. A pedestal-mounted wireless interface with a commercial WiFi chipset has been used in an unconstrained non-human primate [47]. While similar to the BWD in terms of signal sampling and channel count, the package was relatively large, had high power consumption and incorporated a cooling fan. A subsequent commercial device modified from our prototype, Cereplex-W (Blackrock Microsystems) prioritized slightly lower noise, 16-bit 30kS/s sampling, error correction in the wireless link, and more powerful transmission – with the tradeoff of requiring much higher power consumption than the BWD and therefore much shorter operation per battery charge.

Other broadband wireless systems for multielectrode research have been demonstrated in freely moving primates [48]–[53] and a growing number of commercial alternatives support wireless intracortical recording for animal research. To our knowledge none are yet approved for human use, but several designs have a level of integration and other design tradeoffs that could be consistent with future translation. For any of these systems, engineering and translational challenges toward chronic human use include pairing the wireless components to intracranial electrodes suitable for long-term implantation in people, a small and lightweight package, high data rates (and/or on-board processing) across many electrodes, long run time (low power consumption), successful safety testing and completion of regulatory activities.

### B. Comparison to Other Wireless Implanted BCIs

An early wireless broadband BCI demonstrated the ability for an ALS patient to toggle a binary signal from a single intracortical electrode [54]. One early ECoG closed-loop system with 32 subdural electrodes enabled an individual with tetraplegia [55] to move a cursor to targets by associating four different visualized movements with four cursor directions. Although neither wireless nor chronic, this system demonstrated moderate multidimensional cursor control based on decoded intracranial signals. A novel approach to intracranial recording, via electrodes attached to an endovascular stent [56], is now in clinical feasibility trials.Such systems, while very low bandwidth, may represent an effective solution for some individuals with severe disability.

Recently, deep brain stimulators (DBS) have served as a convenient, clinically accepted platform upon which fully implanted wireless BCIs have been developed [57]–[59]. A wireless BCI, configured from a bi-directional fully implanted DBS device (Activa PC+S, Medtronic, Inc.) was used to sample field potential data (0.8kS/s) from a bipolar pair of chronic subdural ECoG electrodes in a person with ALS [58]. The wireless link from the subclavicular transmitter (11.7 kbps) was more than 1,000x lower bandwidth than one BWD; nevertheless, this enabled the individual to use the BCI to make click selections in a commercial scanning letter interface. The research participant spelled 44 prompted words at a mean rate of 1.15 correct characters per minute (without letter prediction). Despite the slow communication rate, the participant reported satisfaction with the system. In another study, the PC+S system was used for clinical stimulation and ECoG recording of motor-related potentials from five patients with Parkinson’s Disease over the span of a year [59]. Up to 9 minutes of data from two subdural sites were recorded at a time and then downloaded for offline analysis. Although no closed-loop study was attempted, changes in beta and gamma bands were observed during movement, possibly relevant to a future closed-loop BCI implementation. The W-HERBS 128-channel ECoG system with 1kS/s sampling per channel has been developed over many years but not yet tested *in-vivo* [60]. An ECoG system specifically designed for translation, the WIMAGINE® ECoG neural interface (Clinatec, Grenoble, FR), was recently demonstrated in an individual with spinal cord injury [61]. Two devices were placed over bilateral motor cortical areas for chronic epidural recording. The wireless data link supported 586 samples/second x 32 contacts using external antennas held near the implants by a custom helmet. Using the BCI to activate a neural on/off switch, the participant was able to initiate walking movements of an avatar and an exoskeleton. The subject also commanded continuous control of bilateral limb movements of the exoskeleton and was able to steer a powered wheelchair. We have not yet evaluated the current wireless system relative to intracortical cabled control of robotic, prosthetic, or functional electrical stimulation applications [1]– [3], [11], [12]. However, the higher electrode count and 100x data rates demonstrated here provide the potential for more dexterous low-latency wireless motor prostheses. Whether ECoG BCIs can approach the communication performance demonstrated here remains to be determined.

### C. Leveraging Broadband Wireless Recording

While the appropriate recording fidelity for stable, high-performance intracortical BCIs in people remains an active area of research, recent advances in high-performance iBCIs have leveraged broadband recording by decoding discrete spike rates combined with power in high-frequency local field potentials (HF-LFPs) up to 450 Hz [7], [12], [16] or 5 kHz [4], [27]. This “hybrid” decoding has contributed to stable high performance iBCI cursor control [4], [7] and rapid decoder calibration [27]. This and other iBCI studies have incorporated common average referencing computed across a subset of electrodes at high sample rate (e.g., 500 Hz [16]; 15 kHz [4], [7], [27]) to reduce noise prior to band pass filtering that could otherwise alias noise into the features computed for decoding. This technique computes a common signal from a subset of channels in real-time and applies it to all electrodes prior to computing neural features for decoding. Here, threshold crossing spike rates were computed after first applying a non-causal (bidirectional two-pass) bandpass filter with a 5 kHz upper cutoff frequency that has been shown to measurably improve decoding and BCI cursor performance relative to simple causal thresholding [29]. While we have not yet rigorously established a precise bandwidth that is necessary or sufficient for iBCI performance and stability for people with tetraplegia, these current practices provide a possible reference point for future optimized wireless BCIs.

### D. Future Work

One advantage of recent ECoG or stent-based BCIs relative to the external wireless system in the current study is that they are fully implanted with no percutaneous connections. A fully-implantable version of the BWD [25], [26], modified with a titanium package and inductive power, has been tested in animals and is being readied for regulatory testing prior to potential deployment in human clinical trials. To that end, the wireless device used in this study incorporates major elements of the fully implanted design and represents an externally-mounted prototype of those core technologies. We have also demonstrated a prototype battery-powered mobile embedded decoding system [62]–[64] that integrates with both the external and fully implanted transmitters and is readily interfaced with other future electrode technologies with hundreds or thousands of contacts.

When integrated with stimulation capabilities, human wireless intracortical systems will offer new opportunities beyond iBCI [65] in neural sensing and closed-loop neuromodulation, including the treatment of seizures and neuropsychiatric disease [66]–[68]. Limitations of this current generation BWD and its implantable sibling are their unidirectional communication; neither stimulation nor impedance testing are currently possible. ASIC chips are being actively developed to introduce stimulation and impedance spectroscopy to future generations of this family of wireless devices.

Looking to the future, features of neural activity beyond spiking (e.g., high-frequency field potential oscillations, field-field phase coherence, spike-field relationships) remain to be fully explored in people and could one day be leveraged to enhance iBCI capabilities, performance and robustness. High resolution intracortical recording – currently only feasible as part of clinical research endeavors – will facilitate continued investigation of aspects of neural activity that may be clinically useful. We anticipate significant advances and paradigm shifts in neural signal processing, decoding algorithms, and control frameworks that will advance neural decoding for effective iBCI and neuromodulation applications. When, how and if different groups (academic, industry, etc.) should diverge in their efforts to develop better and clinically viable iBCI systems is a discussion central to iBCIs’ successful translation.

## V. Conclusion

This study reports important progress in the clinical translation of an iBCI toward a future assistive medical device for individuals with paralysis. We demonstrate the first high-resolution broadband recording from multiple implanted microelectrode arrays using a wireless iBCI in human subjects. Results with two individuals with tetraplegia demonstrate the viability of a wireless iBCI for real-time control of a point-and-select interface. Characteristics of the BWD recordings were highly comparable to those of wired recordings. This broadband wireless system also enables ongoing fundamental research into cortical processing during every day human behavior to inform future neuroscience and BCI advancements. This work overcomes former barriers to in-home mobile independent use of a promising assistive technology to restore communication and digital access for individuals with severe speech and/or motor impairments.

## Data Availability

The sharing of the human neural data is restricted due to the potential sensitivity of this data. Data may be made available upon request to the authors (J.D.S. or L.R.H.). To respect the participants’ expectation of privacy, a legal agreement between the researcher's institution and the BrainGate consortium would need to be set up to facilitate any sharing of these datasets.

## Acknowledgment

We thank participants T10, T5 and their families and caregivers; K. Barnabe, M. Bowker, B. Travers and Y. Mironovas.

## Conflict of Interest

The MGH Translational Research Center has a clinical research support agreement with Neuralink Corp., Paradromics, and Synchron, for which LRH provides consultative input. KVS is a consultant to Neuralink Corp., and on the Scientific Advisory Boards of CTRL-Labs, Inscopix Inc., Heal Inc. and Mind X. JMH is a consultant for Neuralink Corp., Proteus Biomedical and Boston Scientific, and serves on the Medical Advisory Boards of Enspire DBS and Circuit Therapeutics.

## Disclaimer

The content is solely the responsibility of the authors and does not necessarily represent the official views of the NIH, the Department of Veterans Affairs or the US Government.

## Caution

Investigational Device. Limited by Federal Law to Investigational Use.

## Notes

### Competing Interest Statement

The Massachusetts General Hospital (MGH) Translational Research Center has a clinical research support agreement with Neuralink, Paradromics, and Synchron, for which LRH provides consultative input. KVS is a consultant to Neuralink Inc., and on the Scientific Advisory Boards of CTRL-Labs, Inscopix Inc., Heal Inc. and Mind X. JMH is a consultant for Neuralink Corp., Proteus Biomedical and Boston Scientific, and serves on the Medical Advisory Boards of Enspire DBS and Circuit Therapeutics.

### Clinical Trial

NCT00912041

### Funding Statement

This work was supported in part by NIH-NINDS UH2NS095548; the Office of Research and Development, Rehab. R&D Service, Department of Veterans Affairs (N9228C, N2864C, A2295R, B6453R, P1155R); NIH-NIDCD R01DC009899 and R01DC014034; NIH-NBIB R01EB007401; The Executive Committee on Research (ECOR) of Massachusetts General Hospital; MGH-Deane Institute; DARPA REPAIR; Wu Tsai Neurosciences Institute at Stanford; Larry and Pamela Garlick; Samuel and Betsy Reeves; The Howard Hughes Medical Institute; Conquer Paralysis Now 004698.

